# Capturing the SARS-CoV-2 infection pyramid within the municipality of Rotterdam using longitudinal sewage surveillance

**DOI:** 10.1101/2022.06.27.22276938

**Authors:** Miranda de Graaf, Jeroen Langeveld, Johan Post, Christian Carrizosa, Eelco Franz, Ray.W. Izquierdo-Lara, Goffe Elsinga, Leo Heijnen, Frederic Been, Janko van Beek, Remy Schilperoort, Rianne Vriend, Ewout Fanoy, Evelien I.T. de Schepper, Marion P.G. Koopmans, Gertjan Medema

**Author notes:** These authors contributed equally to this article (Miranda de Graaf, Jeroen Langeveld, Johan Post). These senior authors contributed equally to this article (Marion P.G. Koopmans, Gertjan Medema).

## Abstract

**Background:** Despite high vaccination rates in the Netherlands, severe acute respiratory syndrome coronavirus 2 (SARS-CoV-2) continues to circulate. Longitudinal sewage monitoring was implemented along with the notification of cases as two parts of the surveillance pyramid to validate the use of sewage surveillance for monitoring SARS-CoV-2, as an early warning tool, and to measure the effect of interventions.

**Methods:** Sewage samples were collected from nine neighborhoods from September 2020 to November 2021, and compared with reported cases. Comparative analysis and modeling were performed to understand the correlation between wastewater and case trends.

**Findings:** Using high resolution sampling, normalization of wastewater SARS-CoV-2 concentrations and ‘normalization’ of reported positive tests for testing delay and intensity, the incidence of reported positive tests could be modeled based on sewage data, and trends in both surveillance systems coincided. The high collinearity implied that high levels of viral shedding around the onset of disease largely determines SARS-CoV-2 levels in wastewater and the observed relation was independent of SARS-CoV-2 variants and vaccination levels.

**Interpretation:** Wastewater surveillance can accurately display SARS-CoV-2 dynamics for small and large locations, and is sensitive enough to measure small variations in the number of infected individuals within or between neighborhoods. With the transition to a post-acute phase of the pandemic, continued sewage surveillance can help to keep sight on reemergence, but continued “pyramid” validation studies are needed to assess the predictive value of sewage surveillance with new variants.

**Funding:** Horizon H2020, Adessium Foundation, STOWA, TKI, Ministry of Health, Welfare and Sport

## Introduction

The coronavirus disease 2019 pandemic is a major challenge for society, and to control outbreaks the largest global testing program in history was started ^1^. For most pathogens, there are differences in the severity of disease between infected individuals, with a small group experiencing symptoms that are so severe that hospital admission is warranted, while a much larger group will experience mild symptoms or undergo asymptomatic infection ^2,3^. Since only people that seek medical care will be diagnosed, an outbreak can occur relatively unnoticed for some time ^4,5^. SARS-CoV-2 is an example of a viral pathogen for which the disease pyramid is heavily skewed towards mild and asymptomatic cases ^5^.

Over the course of the pandemic in the Netherlands, the testing capacity testing was increased, so that also those with mild symptoms or that were in close contact to SARS-CoV-2 positive individuals could be tested. In addition, as the pandemic progressed a negative test was needed for travel and events, and self-testing became more frequent. As a result, SARS-CoV-2 testing varied between locations and over time.

Wastewater can be used for population surveillance of viruses that may spread by persons with mild or asymptomatic disease, and has been used for common endemic pathogens, antimicrobial resistance ^6^ and poliovirus ^7,8^. SARS-CoV-2 can be detected in domestic wastewater and levels of SARS-CoV-2 in wastewater correlate with reported cases and hospitalizations ^9,10^. In the Netherlands, during the spring of 2020 SARS-CoV-2 was already detectable in city wastewater six days before the first patients were diagnosed ^11^. In addition, next generation sequencing (NGS) and digital droplet PCR (ddPCR) were used to test for the presence of SARS-CoV-2 variants of concern (VOC) and to unravel the genetic characteristics of SARS-CoV-2 viruses present in sewage ^12,13^.

In other countries wastewater surveillance also outperformed case reports as a tool to monitor SARS-CoV-2 circulation, where the detection of SARS-CoV-2 in wastewater preceded case reports with 3 to 14 days ^9,10,14,15^. The added value of sewage surveillance, depends on multiple factors, such as testing access and policy, healthcare access, demographics, and compliance to policies at community level. Therefore, we conducted a high-resolution surveillance project in the Rotterdam-Rijnmond area, to assess how the dynamics of the number of clinical cases within a neighborhood are reflected in viral loads in sewage and which factors affect this correlation. We further investigated the potential of sewage surveillance to monitor SARS-CoV-2 trends, as early warning tool, and to measure the effect of interventions. For this, we designed a study to collect the following data from different layers of the surveillance pyramid: 1) high resolution sewage sampling in 9 neighborhoods of different sizes and socioeconomic status within Rotterdam, 2) notification data based on clinical testing, with number of notifications, dates of probable day of onset of disease, and dates of test result from the national notification database for community cases in the same neighborhoods (OSIRIS), 3) data on total number of tests performed in the testing lanes of the regional Public Health Service (PHS). All data sources were matched per neighborhood based on zip codes. During this study, sewage surveillance was done in real-time, and shared with municipal public health experts to assess usefulness of this type of data in decision making on SARS-CoV-2 mitigation measures for Rotterdam.

## Methods

### Sample preparation

Automated samplers were installed during the summer of 2020, sampling of the Bergschenhoek area started in December 2020 upon an outbreak of the alpha VOC. Wastewater specimens were collected three times per week as 24h flow-dependent composite samples and processed as previously described ^11,16^. Samples were screened by RT-qPCR with primer and probe sets targeting the SARS-CoV-2 N2 and E-gene and CrAssphage.

### Normalization sewage data

The sewage in Rotterdam consists of domestic wastewater from households, industrial wastewater, extraneous waters, such as infiltrating groundwater or inflowing surface water, and runoff. All non-domestic wastewater flows can dilute SARS-CoV-2 levels and vary strongly in time and per catchment, therefore normalization was applied to enable comparison over time and between catchments ^16^. The samples were normalized based on the quotient of the measured daily volumes of sewage and the expected amount of domestic wastewater (the average volume of domestic wastewater produced per person and day multiplied by the population per sewer district). This normalization was verified using the conductivity of the sewage and levels of crAssphage (an indicator of human fecal contamination) and erroneous data points were removed ^16^. The total inflow at WWTP Dokhaven was calculated as the mean flow weighted mean of the four larger catchment areas.

### Data preparation and model exploration

Dynamic time warping was used to explore the lag-times between the datasets ^17^. In order to account for the delay in testing, the date of first illness was assigned to the test results using the average number of days between first day of illness and testing on each date.

The normalised sewer data and the time-shifted test data were used to derive a first linear regression model. To account for variations in testing intensity an updated model was developed by (1) fitting a simple linear regression model with all explanatory variables, (2) adding a random component, if necessary, (3) select the appropriate variance structure, (4) select an appropriate correlation structure, (5) derive the optimal fixed structure with explanatory variables. Incidence served as response variable. The normalised sewage data served as an explanatory variable and the total number of tests performed per day per 100,000 people was added in this model. The model with a fixed variance was outperformed by a model with different variances per area to account for area size dependent noise in the response variable and yielded a lower Akaike Information Criterion (AIC) score. An autoregressive component of the order 1 was added due to the independence assumption; residuals from successive time points showed signs of correlation which may lead to unreliable estimates of explanatory variable variances. Noise in the model estimates was reduced by adding a thin plate regression spline as a smoothing function on the date. Cross validation was applied to optimize the amount of smoothing and prevent overfitting. This resulted in the following formula.

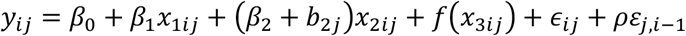

Where *y*_*i,j*_ is the response variable, the reported number of positive tests per 100,000 on day *i* in catchment *j*, time-shifted towards the first day of illness. *β* are the fixed effect coefficients derived for the intercept (*β*_0_) and the explanatory variables (*β*_1_, *β*_2_). The first explanatory variable is the normalized SARS-CoV-2 concentrations in wastewater *x*_1_. The reported total number of tests per 100,000 *x*_2_ represents the testing intensity and served as a measure of the willingness to test. As this relation may differ per location, the random slope *b*_2*j*_ was added. *f(x*_*3ij*_) is a smoothing function represented by cubic regression smoothing splines used to reduce noise in successive modelled values of the time series. Both the random slope *b*_2*j*_ and the error term *ϵ*_*ij*_ are assumed to be normally distributed as 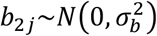 and 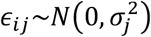, where the latter has a standard deviation that varies per catchment. *ρε*_*j,i*−1;_ refers to the error of a first-order autoregressive AR(1) component that is accounting for autocorrelation in the time series. The model was implemented in the Linear and Nonlinear Mixed Effects Models and generalised cross-validation package in R Version 4.0.5 ^18-20^

### PHS testing lanes

SARS-CoV-2 test results were extracted from the database based on zipcode of the residential address. Subjects with or without self-reported symptoms were tested by RT-PCR ^21^. This study was approved by the Erasmus MC Medical Ethical Committee (MEC-2020-0617).

## Results

### Selection of areas

In total, 9 catchment areas representing between 6,500 and 138,280 inhabitants were included in this study. We first selected four neighborhoods for which the coverage area of general practices overlapped well with the sewer catchment areas: Katendrecht, Ommoord, Pretorialaan and Rozenburg. In addition, we selected four larger catchment areas: influent (INF)2 Everlo-Waalhaven, INF3 Pretorialaan-Zuidplein, INF4 Wolphaertsbocht and INF5-6 Heemraadsplein for which we could collect sewage samples at wastewater treatment plant (WWTP) Dokhaven (Figure1A). The sewer systems of Katendrecht, Pretorialaan and INF3 Pretorialaan-Zuidplein form a cascade were upstream catchments discharge into larger downstream catchments. We later included Bergschenhoek during a large-scale testing effort. Sampling occurred between September 2020 and November 2021.

**Figure 1.**
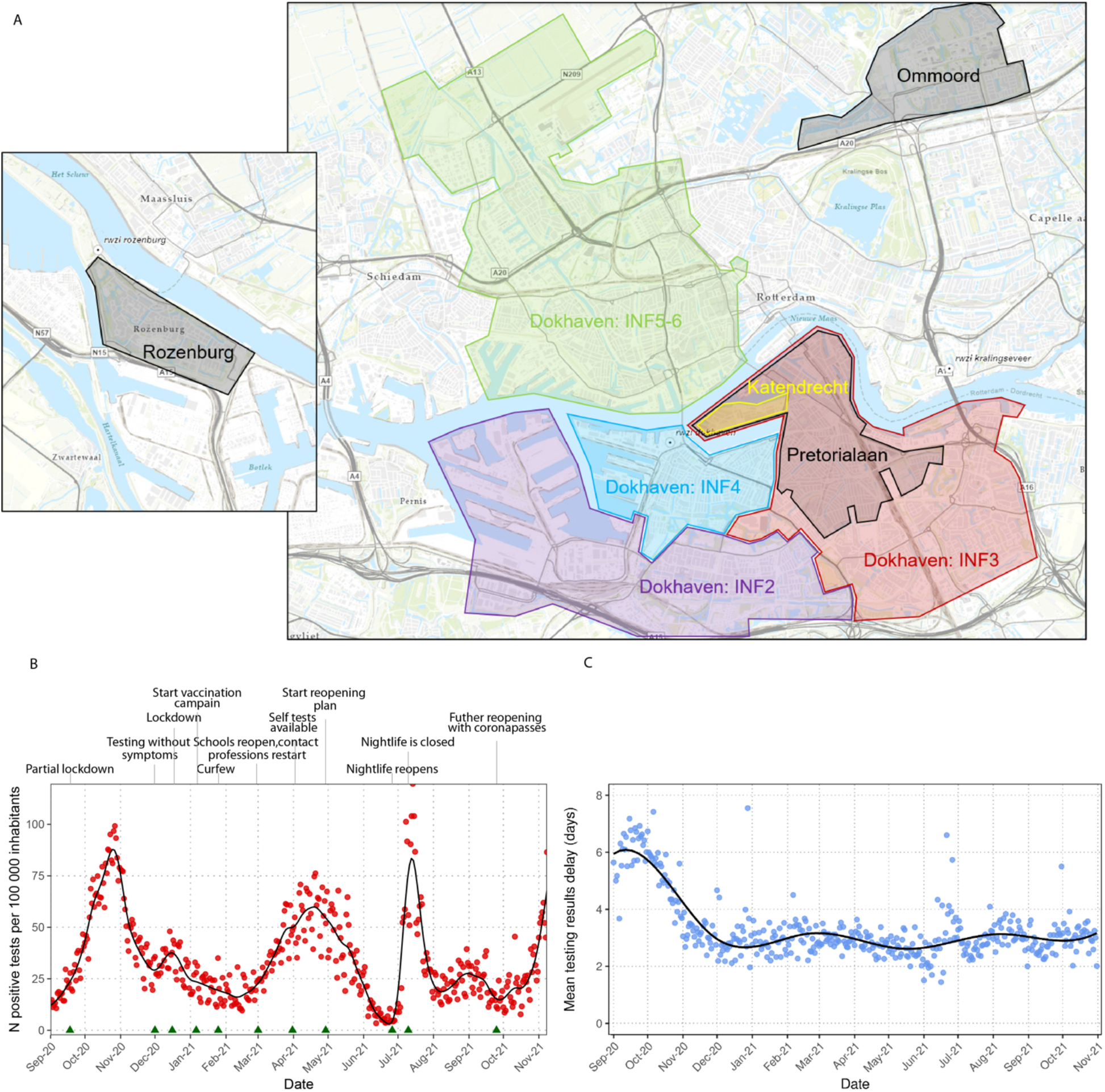
A) Research areas within Rotterdam Rijnmond. Sample collection occurred at wastewater pumping stations Ommoord, Katendrecht, Pretorialaan and Bergschenhoek and wastewater treatment plants (WWTP) Rozenburg and Dokhaven. At WWTP Dokhaven samples were collected from 4 sewer pipes that service the catchment areas INF2 Everlo-Waalhaven, INF3 Pretorialaan-Zuidplein, 4 Wolphaertsbocht and INF5/6 Heemraadsplein. B) SARS-COV-2 positive tests per 100,000 inhabitants for all selected areas within Rotterdam-Rijnmond (excluding Bergschenhoek) and a selection of the implemented lock-down measures during this period (green triangles) for the period September 2020 till October 2021. C) Time from onset of symptoms to test results in total Dokhaven. Cubic regression splines were used to produce the smoothed curves.

### Trends in SARS-CoV-2 notifications per catchment area

Positive cases per 100,000 inhabitants over time were plotted for Rotterdam-Rijnmond (Figure 1B) and per area (Figure 2) and showed peaks in October 2020, December 2020, April 2021, July 2021, and a rise in infections in October 2021. In September 2020, the time from onset of symptoms to reporting a positive test took an average of 4-6 days and this gradually reduced to an average of 3 days after ramping up of test capacity (Figure 1C).

**Figure 2.**
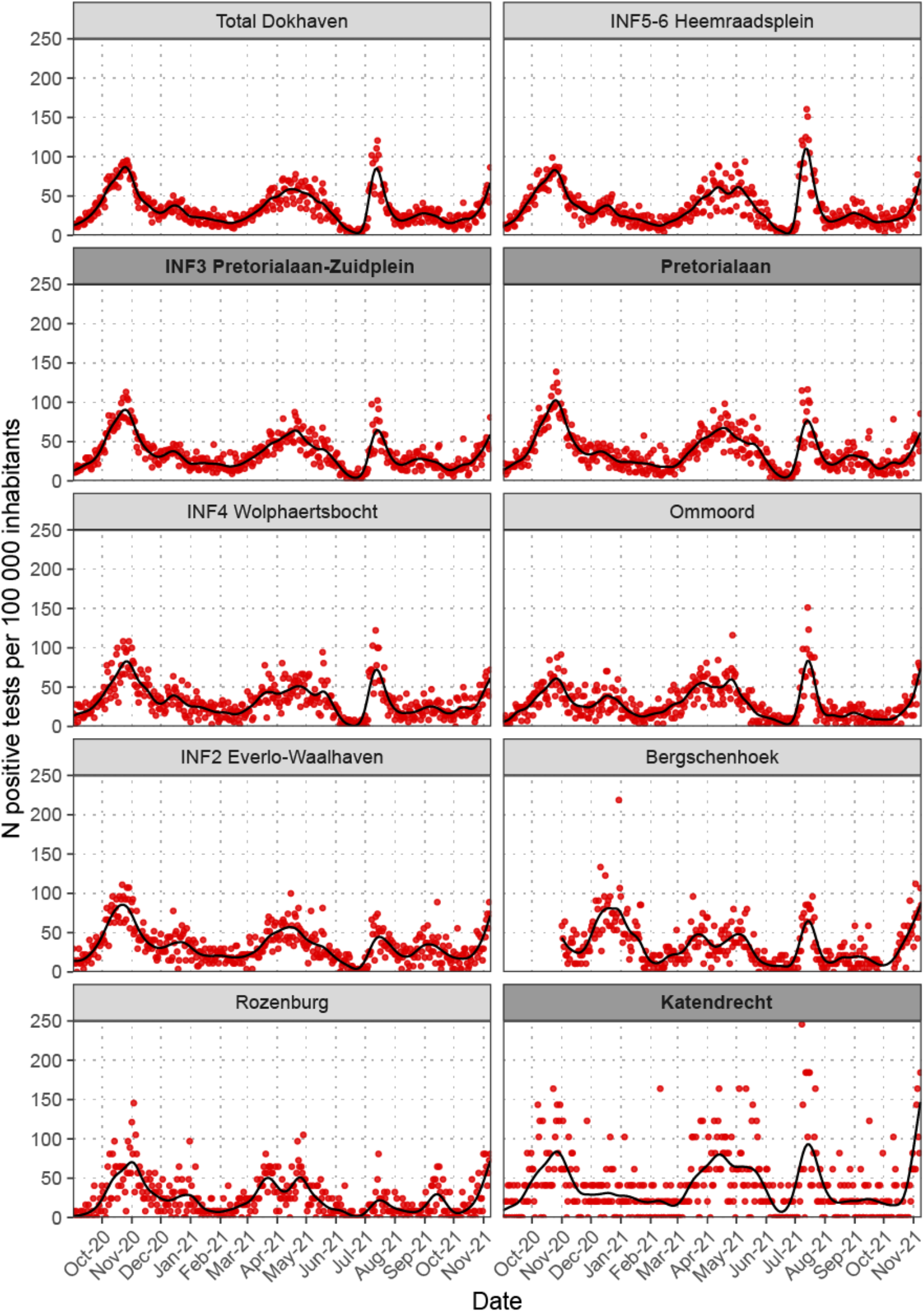
Daily reported SARS-COV-2 positive tests per 100,000 inhabitants for all ten areas per sampling date. The areas are ordered based on population size. Katendrecht to Pretorialaan and INF3 (bold) overlap but are increasing in size. Cubic regression splines were used to produce the smoothed curves.

Overall, all nine catchment areas show similar trends in incidence of reported SARS-CoV-2 cases over time (Figure 2). For the (small) catchment areas, Katendrecht, Ommoord and Rozenburg the signal was noisier compared to the other catchment areas. This was most likely related to the size of the catchment population, as the noise was reduced comparing the cascading dataset of Katendrecht, Pretorialaan and INF3 Pretorialaan-Zuidplein. While overall trends were similar, the communities in the perimeter of the city, “Rozenburg” and “Ommoord”, on average had lower levels of reported positive tests. Also, the rise in infections after reopening of the Dutch nightlife in July 2021, leading to a massive spike in cases among young adults, was not observed for Rozenburg, which has an older population, while this increase was largest for the inner-city areas “Katendrecht” and “center of Rotterdam” (INF5-6). The total number of tests varied over time (Supplemental Figure 1), related to changes in testing policy, behavior, and availability.

### Validation of sewage surveillance by comparison with case notification data

The measured concentrations of SARS-CoV-2 in sewage were normalized for the flow of non-domestic wastewater ^16^. SARS-CoV-2 levels in wastewater were highest in October 2020 for all areas but Rozenburg where it was highest in April 2021. SARS-CoV-2 levels were relatively low in Rozenburg and Bergschenhoek.

Initial plotting and calculation of the autocorrelation between the reported SARS-CoV-2 cases and viral loads in wastewater showed that in the period September - December 2020 the highest correlation was observed for a time lag of 6 days between an increase or decrease in the sewer and reported SARS-CoV-2 cases (Supplemental Figure 2). However, with increasing testing capacity, this time delay became shorter (Figure 1). After correcting the reported case data for the time of symptom onset, the incidence, and the concentrations of SARS-CoV-2 in sewage were highly collinear, especially during the raise of the October 2020 wave (Figure 3). This high collinearity observed when correcting for day of illness onset implies that high viral shedding in the sewer around the onset day of new cases, largely determines the SARS-CoV-2 concentration in wastewater, even though fecal shedding can last for more than two weeks ^22-25^. There were two discrepancies: the peak in incidence when the nightlife reopened (July 2021), and a peak in December 2020, possibly reflecting increased testing ahead of seasonal festivities (Figure 3, supplemental Figure 3).

**Figure 3.**
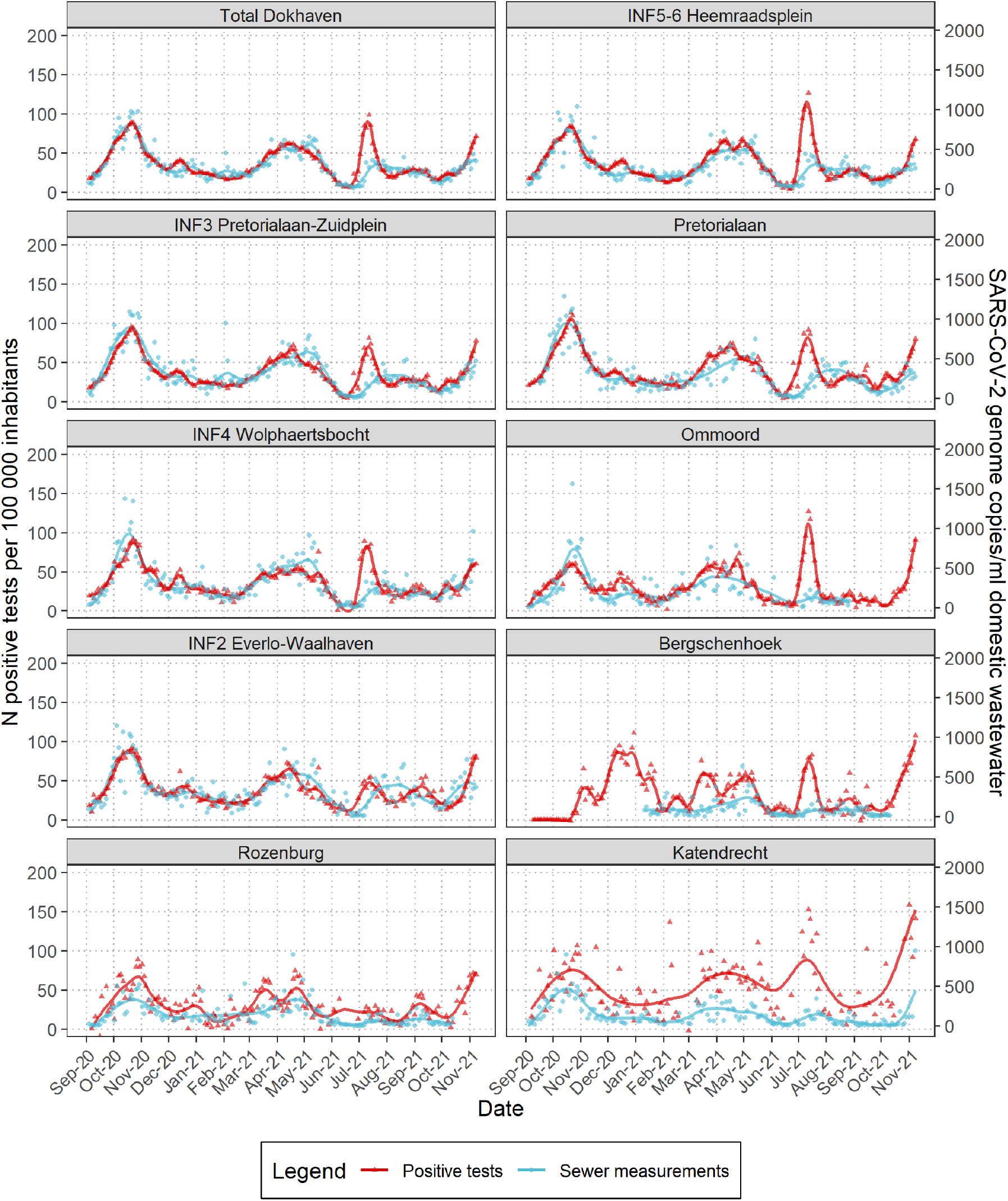
Levels of normalized SARS-CoV-2 genome copies per ml of domestic wastewater in Rotterdam-Rijnmond (blue) and the number of SARS-CoV-2 positive tests per 100,000 corrected for first day of illness (red). The total inflow at WWTP Dokhaven is calculated as the flow averaged mean of the four larger catchment areas. The areas are ordered based on population size. Cubic regression splines were used to produce the smoothed curves.

To investigate these differences, we modeled the expected incidence of reported positive cases by first day of illness based on the normalized SARS-CoV-2 concentrations in wastewater (Figure 4). The model contained a location-specific variance and an autoregressive component, and the best fit was obtained when a random slope for the “total number of tests per population” was added, indicating that changes in testing intensity affected the correlation between sewage and incidence trends (supplementary Figure 4), and that the total number of tests per population should be included in the model. The December 2020 peak of reported cases that was not evident from sewage surveillance, could now be modeled based on a small increase in sewage and a large increase in clinical tests. However, at the height of the outbreak in July 2021 values predicted by sewage were still significantly lower than the reported incidence of SARS-CoV-2 positive individuals, after this outbreak the correlation between wastewater and positive cases was restored.

**Figure 4.**
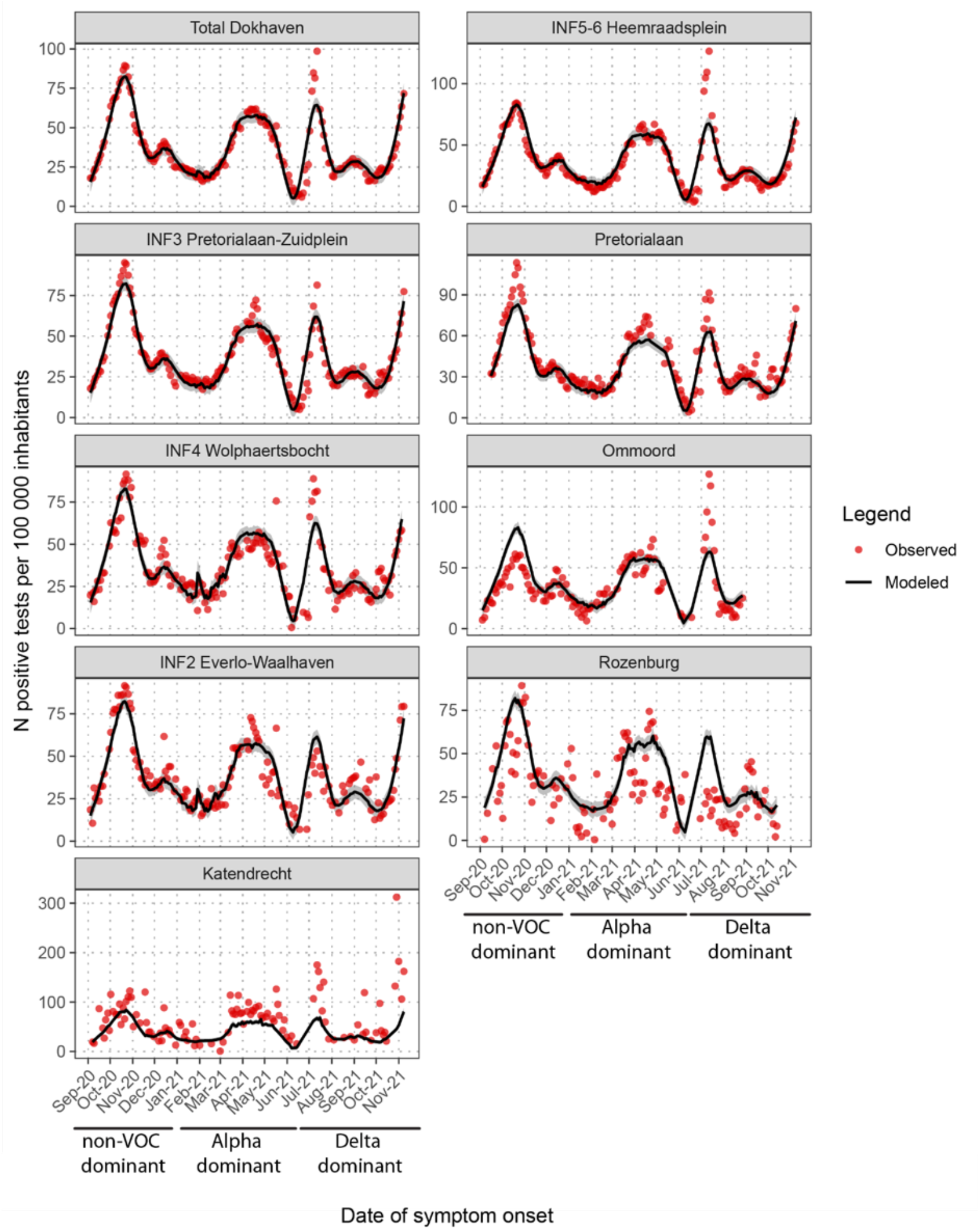
Modeling of the expected incidence of reported cases based on wastewater. Reported SARS-CoV-2 positive tests over time were shown in red with the predicted incidence of reported SARS-CoV-2 positive tests based on the normalized SARS-CoV-2 concentration in sewage for each of the areas. The model contained a location-specific variance and the incidence data was corrected for testing delay and testing intensity.

The correlation was high (R^2^ > 0.75) in areas with larger population but weak for areas with a population <25,000 (R^2^ < 0.37). Plotting the correlation as a function of the population size (Figure 5A) indicates that this correlation was predominantly limited by noise in incidence data and not in the sewage surveillance data. In agreement with statistical theory, this noise reduced with an increasing sample size and was roughly proportional to the square root of *n*.

**Figure 5.**
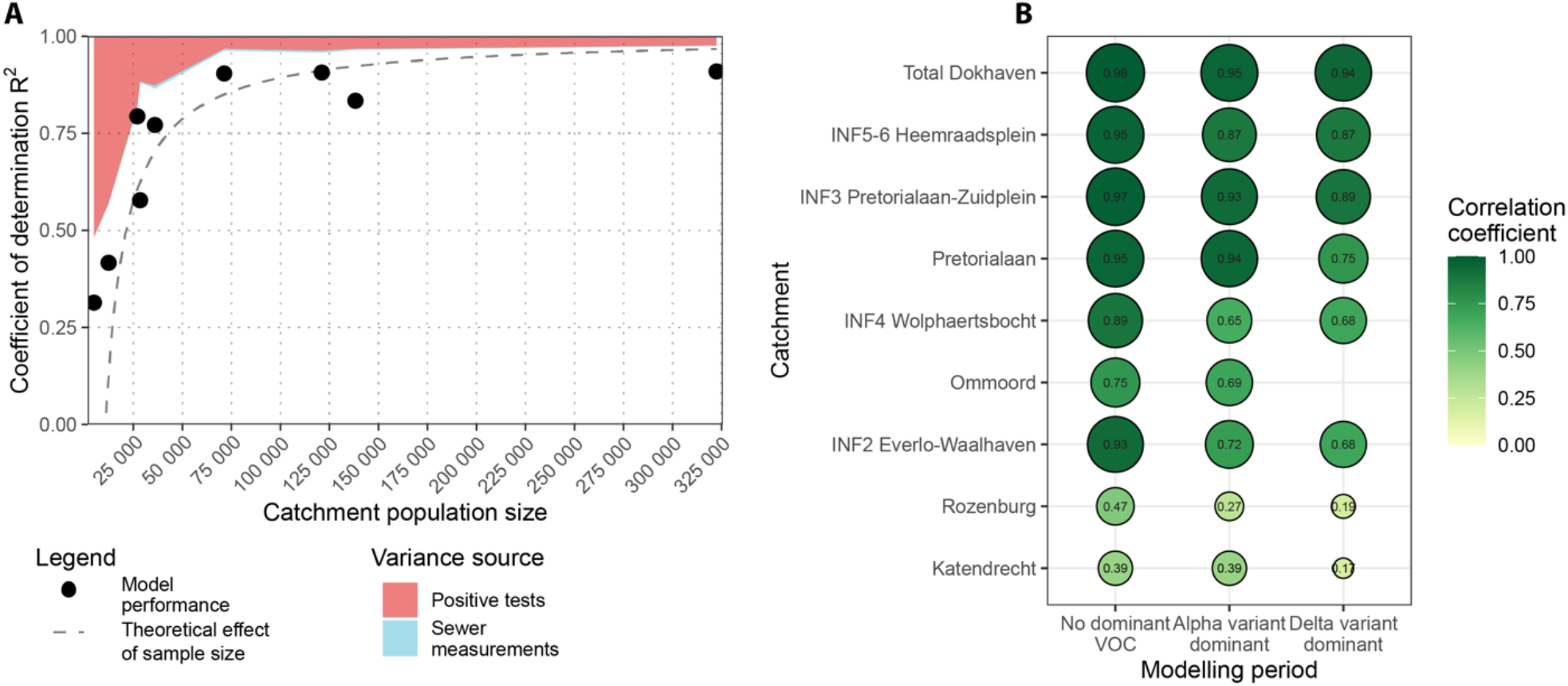
A) Goodness-of-fit test for the applied additive mixed model for each of the nine locations. The population size is given on the X-axis, model performance expressed in R^2^ on the Y-axis. Duplicate analysis was used to estimate the variance in sewage surveillance data measurements inherent to the testing method. Variance of the reported number of positive tests was estimated using the differences in daily test incidence data after correcting for weekly trends. Theoretical effect is given by the square root of the population size. B) Correlation between the predicted number of reported cases based on sewage and the reported number of positive tests. The areas are ordered based on population size. The predicted number of positive tests for each location and modeling period are calculated using predictive model parameters based on data from period 1 “no dominant variant”. The correlation coefficient is depicted in color and numbers.

During the sampling period, the initial circulating SARS-CoV-2 strains were replaced by the alpha and later by the delta VOC. As differences in viral characteristics could lead to changes in tropism or shedding rates, these replacements could affect the ratio of SARS-CoV-2 levels in wastewater versus the incidence of cases. This was investigated by dividing the data in three different time periods based on VOC circulation; 01-09-2020 till 31-12-2020 (non-VOC), 26-02-2021 till 13-06-2021 (alpha VOC) and 23-07-2021 till 09-11-2021 (delta VOC) ^26^. For the three catchments with the largest population (pop. >120,000) the model parameters established in the first period adequately described the data from the second (alpha) and third (delta) period, resulting in high correlation between wastewater and case notifications for all three periods (Figure 5B). For the intermediate size catchments (pop. 18,000-37,000), the correlation was high in the first period, but lower in the alpha and delta period. For the two smallest catchment areas, the model fit was relatively poor for the first period, due to the variability in both wastewater and clinical surveillance data, and this reduced further in the following periods.

### Sewage surveillance to address public health questions, and to monitor the effect of interventions

In December 2020 there was an outbreak of the alpha VOC in the village Bergschenhoek within municipality of Lansingerland. During this outbreak a large-scale testing effort was initiated for Lansingerland and a sampling cabinet was placed in Bergschenhoek for parallel wastewater monitoring. In Lansingerland between the 11^th^ and 22^nd^ of January 36,534 of 63,338 (58%) residents were tested. For Bergschenhoek 118 positive SARS-CoV-2 tests were reported (629 per 100,000 inhabitants). The average normalized concentration of SARS-CoV-2 in Bergschenhoek wastewater in this period was 66 genome copies (GC)/ml domestic wastewater, indicating each 1/100.000 positive tests would correspond to 66/629 = 0.10 GC/ml of domestic wastewater. The larger catchment areas in Rotterdam (INF3 and INF5/6), where in that same time period only symptomatic individuals were tested (1.9-2.7% of the residents), reported lower incidences of 269 and 293 positives per 100,000 inhabitants. The corresponding average concentrations in sewage in these catchment areas were higher: 151-164 GC/ml domestic wastewater. This would compute to 0.53-0.56 GC/ml per 1/100.000 positive tests, yielding a fivefold difference between large scale population testing (Bergschenhoek) and symptomatic case testing (INF3 and 5/6). With comparable shedding rates between symptomatic and asymptomatic cases ^27^, this would suggest five times higher incidence than reported in INF3 and 5/6.

During the large-scale testing effort levels SARS-CoV-2 in wastewater declined for Bergschenhoek, but increased for INF3 and INF5/6 indicating that this intervention potentially had an effect on SARS-CoV-2 circulation.

## Discussion

Sewage surveillance is now widely used in parallel to testing of suspected cases, and several groups showed the correlation between the number of reported cases and SARS-CoV-2 levels in wastewater ^9-11,14^. Our high-resolution data in well-matched populations along the surveillance pyramid clearly showed that wastewater surveillance was ahead of clinical testing at the start of the second wave in Rotterdam in October 2020, and that this was due to the delay between disease onset, with high virus shedding, and the time a case was reported. Over time, ramping up of clinical testing has reduced this delay and thereby the lead-time of sewage testing for early warning in this setting. Similar results were found for a 14-month-long wastewater surveillance time series in Massachusetts where wastewater data served as an early warning system for the first wave but not during the second, also most likely due to ramping up of testing availability ^28,29^.

While trends based on sewage testing and the incidence of reported cases were highly colinear, there was a clear delineation in July 2021, after the re-opening of the Dutch nightlife and events. Starting June 18, 2021 events were accessible after a negative test and many more (young) people got tested during this period using an antigen test. We hypothesized that a high percentage of individuals got tested in the testing-street after a positive antigen test, while the negative antigen tests do not show up in the testing street results. This was supported by the strong increase in the percentage of positive tests at that time. It also showed that with standard testing a sudden increase in a niche group can quickly be discovered, while this is not possible with sewage surveillance.

We selected multiple neighborhoods and areas within Rotterdam to investigate if SARS-CoV-2 dynamics can vary within a city and if these differences would be evident through wastewater surveillance. Comparison of smaller areas enclosed in larger areas showed similar dynamics. However, in smaller areas relatively few infected individuals can have a large impact on SARS-CoV-2 trends and a weak correlation was observed between reported and predicted SARS-CoV-2 cases for the smallest catchment areas. This was mostly due to noise in the above ground data, and less to noise in the sewage data, meaning that sewage testing even for small catchment areas (serving 6,500 individuals) can be used to identify trends in SARS-CoV-2 circulation. Trends of reported SARS-CoV-2 positives and sewage were similar for each area except Rozenburg and Bergschenhoek, and this was supported by our model, showing that wastewater surveillance can be used to compare neighborhoods.

From our results we deduced that peak shedding of SARS-CoV-2 in feces occurs around the time of symptom onset. This agrees with the viral load in respiratory samples, as the highest levels of virus shedding occurred around onset of symptoms ^23,30,31^. Available human fecal shedding data on SARS-CoV-2 does not show a high initial peak shedding, but most studies do include data around symptom onset ^32^. However, animal studies showed that shedding kinetics in the feces were similar or slightly delayed compared to the respiratory tract ^33-35^.

This study showed the sensitivity of sewage surveillance in relation to the dynamics in case-based surveillance: when the wastewater concentrations were just above the limit of detection in the larger catchments in June 2021, the reported incidence was around 2 per 100.000. During the large-scale surveillance of Bergschenhoek, the ratio between reported incidence and normalized wastewater concentration was around fivefold lower than in the large catchment areas in Rotterdam where only a small percentage of the population was tested, which is indicative of significant undertesting. Not surprisingly, differences in testing behavior affect the relation between sewage and clinical surveillance, highlighting the value of sewage surveillance as being independent of testing behavior. Another potential variable is the emergence of SARS-CoV-2 variants as variant specific phenotypical changes could result in differences in tropism, fecal shedding, age range, symptoms, or vaccine breakthrough infections which could affect the correlation between the incidence of reported cases and wastewater surveillance. For example, 1000-fold higher viral loads were reported for patients infected with the delta VOC compared to the 19A/19B strain ^36^, and 10-fold higher viral loads for the alpha VOC compared to previous non-VOC strains ^12^. Whether these VOC also result in higher fecal shedding is not known. Nevertheless, using the model parameters of the first time period, when non-VOC SARS-CoV-2 were predominant, we were able to accurately predict reported SARS-CoV-2 cases at the time that VOC alpha and beta were predominant, indicating that for wastewater concentrations there was no measurable effect of the emergence of these VOC. During this study SARS-CoV-2 vaccination was rolled out in the Netherlands, but the effect on SARS-CoV-2 levels in wastewater appears to be limited. SARS-CoV-2 breakthrough infections after vaccination could result in lower levels of shedding, but only a 2.8–4.5-fold decrease in respiratory viral load in vaccinated compared to non-vaccinated individuals was reported ^37^.

In conclusion, this work shows that wastewater surveillance can, independently of testing behavior, accurately display SARS-CoV-2 dynamics within a city and is sensitive enough to measure small variations in the number of infected individuals within or between neighborhoods. Sewage surveillance alongside the large-scale testing effort at Lansingerland indicated that many SARS-CoV-2 cases go unreported and that sewage surveillance can be used to monitor the effect of interventions. This is especially relevant in times and locations where the willingness to test is lower, testing facilities have scaled down or when home antigen testing is applied as alternative to clinical tests. As clinical test data provides insight into patient characteristics, sewage data supplements this with insight into the overall infection pressure in a specific catchment area. Sewage surveillance combined with NGS or ddRT-PCR can also provide a rapid insight in the spread of new SARS-CoV-2 variants without the need to analyze thousands of patient samples. We anticipate that wastewater testing should be considered as one of the pillars of future surveillance of (re) emerging viruses.

## Data Availability

All data produced in the present study are available upon reasonable request to the authors

## Acknowledgements

The authors are very grateful for the assistance of Roan Pijnacker, the Water Authorities and WWTP and pumping station operators of Waterschap Hollandse Delta and municipality of Rotterdam, staff of IMD for installing and maintaining the autosamplers and sampling specialists of AQUON. This study was financed by STOWA, TKI Watertechnology in collaboration with Erasmus Foundation, Adessium Foundation, European Union’s Horizon H2020 grant VEO (grant no. 874735), Ministry of Health, Welfare and Sport, H2020 and Waterboards Waterschap Hollandse Delta, Hoogheemraadschap van Delfland and Hoogheemraadschap Schieland en Krimpenerwaard.

## References

1. Mercer TR, Salit M. Testing at scale during the COVID-19 pandemic. Nat Rev Genet 2021; 22(7): 415–26.

2. The Novel Coronavirus Pneumonia Emergency Response Epidemiology T. The Epidemiological Characteristics of an Outbreak of 2019 Novel Coronavirus Diseases (COVID-19) - China, 2020. China CDC Wkly 2020; 2(8): 113–22.

3. Sah P, Fitzpatrick MC, Zimmer CF, et al. Asymptomatic SARS-CoV-2 infection: A systematic review and meta-analysis. Proc Natl Acad Sci U S A 2021; 118(34).

4. Zhu N, Zhang D, Wang W, et al. A Novel Coronavirus from Patients with Pneumonia in China, 2019. N Engl J Med 2020; 382(8): 727–33.

5. Munster VJ, Koopmans M, van Doremalen N, van Riel D, de Wit E. A Novel Coronavirus Emerging in China - Key Questions for Impact Assessment. N Engl J Med 2020; 382(8): 692–4.

6. Hellmer M, Paxeus N, Magnius L, et al. Detection of pathogenic viruses in sewage provided early warnings of hepatitis A virus and norovirus outbreaks. Appl Environ Microbiol 2014; 80(21): 6771–81.

7. Nieuwenhuijse DF, Oude Munnink BB, Phan MVT, et al. Setting a baseline for global urban virome surveillance in sewage. Sci Rep 2020; 10(1): 13748.

8. Hendriksen RS, Munk P, Njage P, et al. Global monitoring of antimicrobial resistance based on metagenomics analyses of urban sewage. Nat Commun 2019; 10(1): 1124.

9. Fernandez-Cassi X, Scheidegger A, Banziger C, et al. Wastewater monitoring outperforms case numbers as a tool to track COVID-19 incidence dynamics when test positivity rates are high. Water Res 2021; 200: 117252.

10. Barrios ME, Diaz SM, Torres C, Costamagna DM, Blanco Fernandez MD, Mbayed VA. Dynamics of SARS-CoV-2 in wastewater in three districts of the Buenos Aires metropolitan region, Argentina, throughout nine months of surveillance: A pilot study. Sci Total Environ 2021; 800: 149578.

11. Medema G, Heijnen L, Elsinga G, Italiaander R, Brouwer A. Presence of SARS-Coronavirus-2 RNA in Sewage and Correlation with Reported COVID-19 Prevalence in the Early Stage of the Epidemic in The Netherlands. Environmental Science & Technology Letters 2020; 7: 511–6.

12. Izquierdo-Lara R, Elsinga G, Heijnen L, et al. Monitoring SARS-CoV-2 Circulation and Diversity through Community Wastewater Sequencing, the Netherlands and Belgium. Emerg Infect Dis 2021; 27(5): 1405–15.

13. Heijnen L, Elsinga G, de Graaf M, Molenkamp R, Koopmans MPG, Medema G. Droplet digital RT-PCR to detect SARS-CoV-2 signature mutations of variants of concern in wastewater. Science of The Total Environment 2021; 799: 149456.

14. Wu F, Xiao A, Zhang J, et al. SARS-CoV-2 RNA concentrations in wastewater foreshadow dynamics and clinical presentation of new COVID-19 cases. Sci Total Environ 2021; 805: 150121.

15. Claro ICM, Cabral AD, Augusto MR, et al. Long-term monitoring of SARS-COV-2 RNA in wastewater in Brazil: A more responsive and economical approach. Water Res 2021; 203: 117534.

16. Langeveld J, Schilperoort R, Heijnen L, et al. Normalisation of SARS-CoV-2 concentrations in wastewater: the use of flow, conductivity and CrAssphage. MedRxiv 2021.

17. Giorgino T. Computing and Visualizing Dynamic Time Warping Alignments in R: The dtw Package. Journal of Statistical Software 2009; 31(7): 1 –24.

18. Wood SN. Fast stable restricted maximum likelihood and marginal likelihood estimation of semiparametric generalized linear models. Journal of the Royal Statistical Society Series B (Statistical Methodology) 2011; 73: 3–36.

19. Dessau RB, Pipper CB. [‘‘R”--project for statistical computing]. Ugeskr Laeger 2008; 170(5): 328–30.

20. Pinheiro J, Bates D, DebRoy S, Sarkar D, Team RC. Linear and Nonlinear Mixed Effects Models. R package version 3.1-155 ed; 2022.

21. Corman VM, Landt O, Kaiser M, et al. Detection of 2019 novel coronavirus (2019-nCoV) by real-time RT-PCR. Euro Surveill 2020; 25(3).

22. Holm-Jacobsen JN, Bundgaard-Nielsen C, Rold LS, et al. The Prevalence and Clinical Implications of Rectal SARS-CoV-2 Shedding in Danish COVID-19 Patients and the General Population. Front Med (Lausanne) 2021; 8: 804804.

23. Cevik M, Tate M, Lloyd O, Maraolo AE, Schafers J, Ho A. SARS-CoV-2, SARS-CoV, and MERS-CoV viral load dynamics, duration of viral shedding, and infectiousness: a systematic review and meta-analysis. Lancet Microbe 2021; 2(1): e13–e22.

24. Zhang Y, Cen M, Hu M, et al. Prevalence and Persistent Shedding of Fecal SARS-CoV-2 RNA in Patients With COVID-19 Infection: A Systematic Review and Meta-analysis. Clin Transl Gastroenterol 2021; 12(4): e00343.

25. Chen Y, Chen L, Deng Q, et al. The presence of SARS-CoV-2 RNA in the feces of COVID-19 patients. J Med Virol 2020; 92(7): 833–40.

26. https://www.coronadashboardrijksoverheid.nl. 2022. https://coronadashboard.rijksoverheid.nl/landelijk/varianten.

27. Lee S, Kim T, Lee E, et al. Clinical Course and Molecular Viral Shedding Among Asymptomatic and Symptomatic Patients With SARS-CoV-2 Infection in a Community Treatment Center in the Republic of Korea. JAMA Intern Med 2020; 180(11): 1447–52.

28. Xiao A, Wu F, Bushman M, et al. Metrics to relate COVID-19 wastewater data to clinical testing dynamics. medRxiv 2021.

29. Aberi P, Arabzadeh R, Insam H, et al. Quest for Optimal Regression Models in SARS-CoV-2 Wastewater Based Epidemiology. Int J Environ Res Public Health 2021; 18(20).

30. Lewis NM, Duca LM, Marcenac P, et al. Characteristics and Timing of Initial Virus Shedding in Severe Acute Respiratory Syndrome Coronavirus 2, Utah, USA. Emerg Infect Dis 2021; 27(2): 352–9.

31. He X, Lau EHY, Wu P, et al. Temporal dynamics in viral shedding and transmissibility of COVID-19. Nat Med 2020; 26(5): 672–5.

32. Medema G, Been F, Heijnen L, Petterson S. Implementation of environmental surveillance for SARS-CoV-2 virus to support public health decisions: Opportunities and challenges. Curr Opin Environ Sci Health 2020; 17: 49–71.

33. Gaudreault NN, Trujillo JD, Carossino M, et al. SARS-CoV-2 infection, disease and transmission in domestic cats. Emerg Microbes Infect 2020; 9(1): 2322–32.

34. Richard M, Kok A, de Meulder D, et al. SARS-CoV-2 is transmitted via contact and via the air between ferrets. Nat Commun 2020; 11(1): 3496.

35. Sia SF, Yan LM, Chin AWH, et al. Pathogenesis and transmission of SARS-CoV-2 in golden hamsters. Nature 2020; 583(7818): 834–8.

36. Li B, Deng A, Li K, et al. Viral infection and transmission in a large, well-traced outbreak caused by the SARS-CoV-2 Delta variant. medRxiv 2021: 2021.07.07.21260122.

37. Levine-Tiefenbrun M, Yelin I, Katz R, et al. Initial report of decreased SARS-CoV-2 viral load after inoculation with the BNT162b2 vaccine. Nat Med 2021; 27(5): 790–2.

