## Supplemental Figure for "Capturing the SARS-CoV-2 infection pyramid within the municipality of Rotterdam using longitudinal sewage surveillance"

**Supplemental Figures**

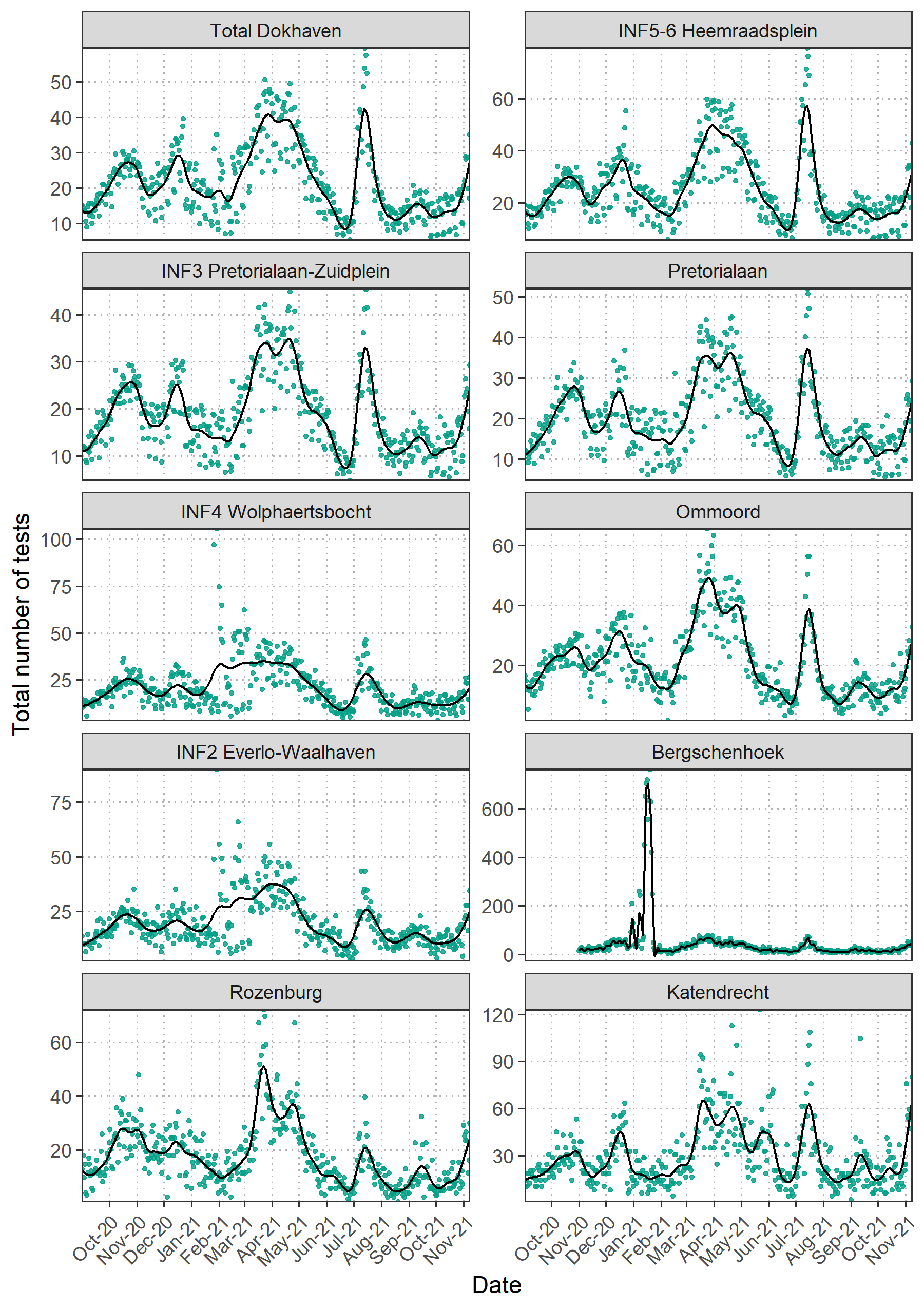

Supplemental Figure 1. Total number test results (N positive +N negative) per 100,000 people per catchment area. Cubic regression splines were used to produce the smoothed curves.

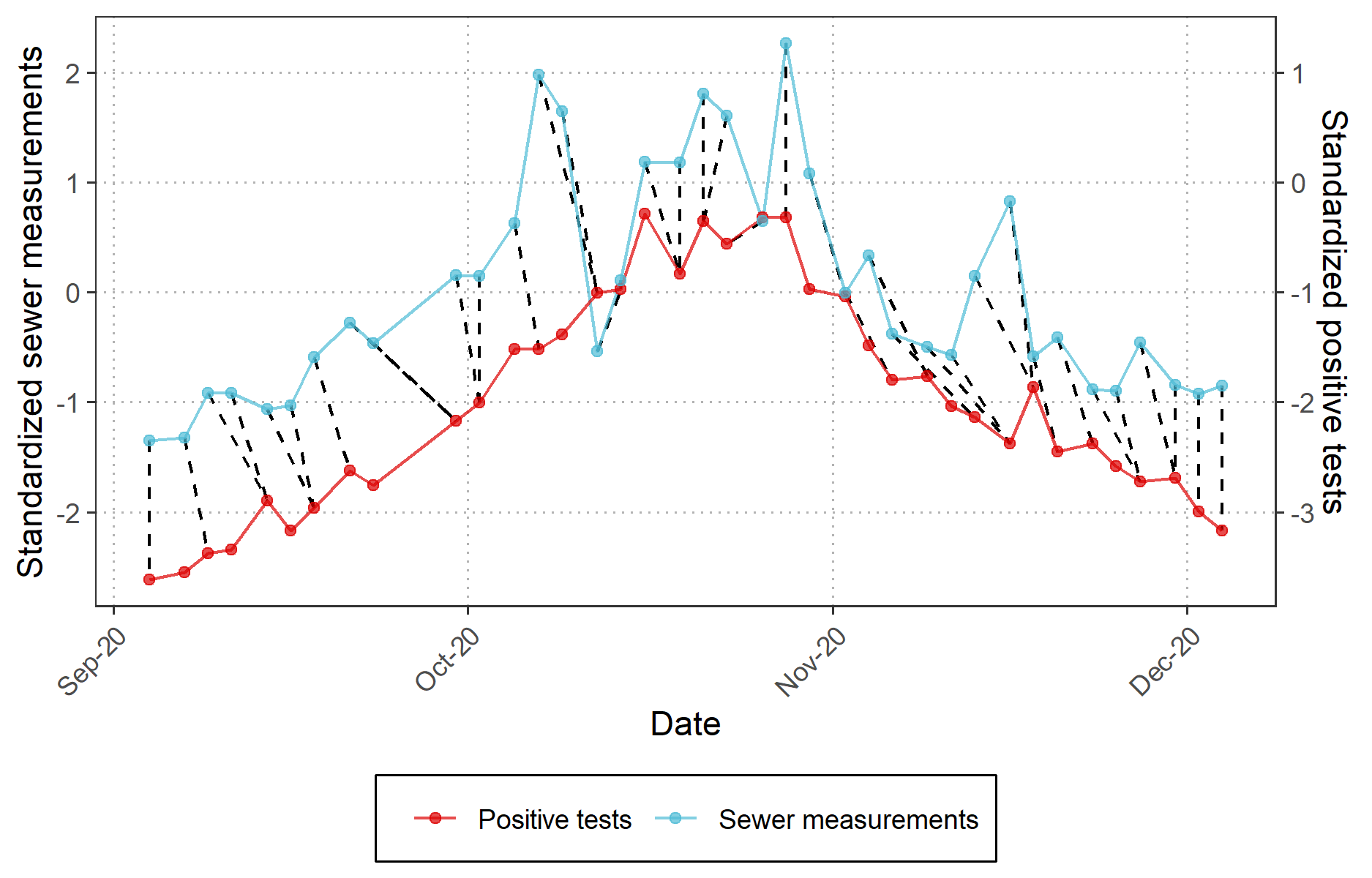

Supplemental Figure 2. Dynamic time warping between the sewer measurements and the SARS-CoV-2 positive tests per 100,000 inhabitants INF5-6 to compare the similarity between the two time series. Dotted lines leaning to the left indicate periods (e.g. end of September) where sewer data is ahead of testing data, while vertical dotted lines indicate a time lag of zero (beginning of December).

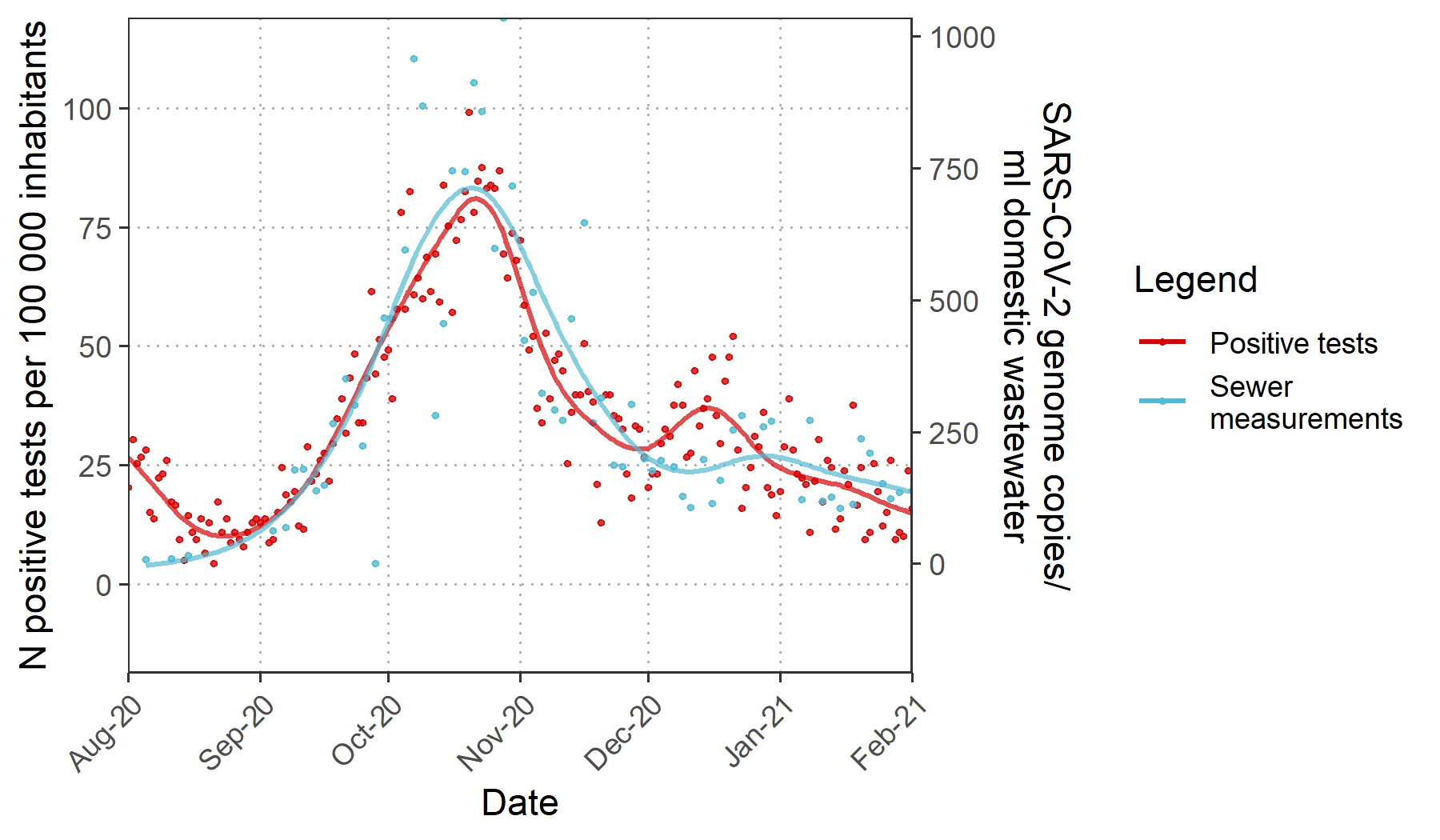

Supplemental Figure 3. Daily SARS-COV-2 positive tests per 100,000 inhabitants for INF5-6 and and levels of normalized SARS-CoV-2 genome copies per ml of household wastewater. Cubic regression splines were used to produce the smoothed curves.

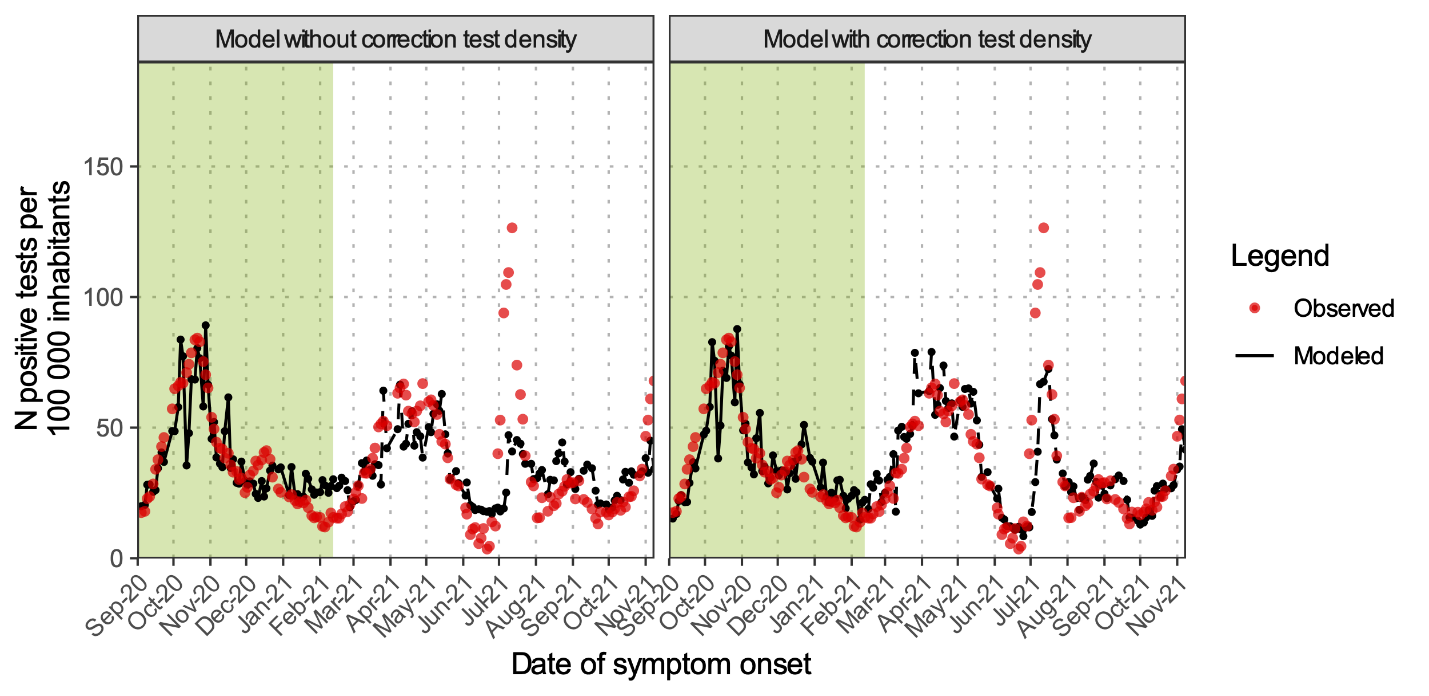

Supplemental Figure 4. Reported SARS-CoV-2 positive tests over time (red) and predicted (black) SARS-CoV-2 positive cases based on sewage from INF5-6. In the left and right panel, the predicted SARS-CoV-2 positive tests were calculated using a model without and with correction for test density, respectively.
